# Early intervention with ColdZyme mouth spray after self-diagnosis of common cold: a randomized, double-blind, placebo-controlled study

**DOI:** 10.1101/2022.06.07.22276075

**Authors:** B. Fredrik Lindberg, Ida Nelson, Jonas Ranstam, Donald K. Riker

## Abstract

**Background:** A glycerol throat spray containing cold-adapted cod trypsin creates a protective barrier and is known to deactivate common cold virus *in vitro* and decrease pharyngeal rhinovirus load after inoculation in humans. Early self-diagnosis and use of the medical device ColdZyme indicate a safe alternative for treatment in naturally occurring common colds.

**Methods:** This was a double-blind, randomized, parallel-group, placebo-controlled study conducted at ten German sites to evaluate the efficacy of the medical device ColdZyme, a glycerol mouth spray containing cold-adapted cod trypsin, for naturally occurring common cold, versus placebo. Adults experiencing a minimum of three common colds during the previous year, but otherwise healthy, were enrolled to begin treatment with the mouth spray or placebo six times daily at first sign of a common cold. Jackson’s symptom scale and the 9-item Wisconsin Upper Respiratory Symptom Survey-21 (WURSS-21) quality of life (QoL) domain and sore throat scale were recorded daily by subjects, as well as any use of allowed rescue treatment. The trial is registered at ClinicalTrials.gov, number NCT03794804.

**Results:** Between January and April 2019, 701 subjects were enrolled and randomly assigned to the ColdZyme group (n=351) or the placebo group (n=350). Of the 701 subjects, 438 (62.5%) subjects developed symptoms typical of common cold, and all 438 started study treatment (n=220 in the ColdZyme group and n=218 in the placebo group).

There were no differences between the groups in primary and major secondary endpoints, however, the assessment using the WURSS-21 QoL domain and Jackson score suggests a slightly faster recovery with ColdZyme, as symptoms and complaints affecting the quality of life were shortened about 1 day. The beneficial effect of ColdZyme was particularly noticeable on the fifth day of the common cold. A positive difference between treatment groups was also seen for the subjects’ assessments of global efficacy of the investigational product, and a robust safety profile for ColdZyme was demonstrated throughout the study.

**Conclusion:** The safety and tolerability of ColdZyme have been confirmed in a larger study population, while establishing evidence of a slightly faster recovery from common cold symptoms.

## Introduction

The common cold is a self-limiting upper respiratory viral infection affecting the individual and also the society in its high costs and lost productivity.^1-2^ The common cold is caused by infection by one of over 200 known respiratory viruses, primarily rhinoviruses, but also coronaviruses, influenza viruses, adenoviruses, parainfluenza viruses, respiratory syncytial viruses and enteroviruses.^3^ Rhinovirus is well adapted to its host, initially overcoming epithelial barriers, interferon release and adaptive immune responses in the nasal and throat mucosa. The diversity of viral pathogens has so far complicated the attempts to find a universal treatment.^4–6^ After infection virus is usually transported from the nose back to the pharynx where infection of the mucosa is first established, followed by anterior spread.^7^ The described migration of the virus finds support in the course of local symptom development, starting with sore scratchy throat and malaise, quickly followed by nasal congestion, rhinorrhea, sneezing and finally cough.^8^

Thus, the first opportunity to halt local virus propagation is during the incubation period. Therefore, a technology has been developed consisting of a hyperosmotic glycerol solution containing cold-adapted trypsin from the Atlantic cod to treat and alleviate the common cold.^9^ When the medical device, ColdZyme, is sprayed orally, the solution forms a barrier on the pharynx that acts osmotically while at the same time interfering with viral attachment and entry. The spray solution has demonstrated broad virucidal activity *in vitro* resulting in a decline of 64-100 % of virus activity when assaying influenza virus, rhinovirus, adenovirus and corona virus, including SARS-CoV-2, using a cytopathic host cell incubation test.^10-12^ In a double-blind study on healthy volunteers inoculated with rhinovirus-16, the barrier solution resulted in a reduced pharyngeal virus load significantly lower than compared to placebo.^13^

The current study was preceded by a single-blind study conducted in the same type of population (i.e. individuals with frequent common colds) to assess whether the study design could adequately evaluate the common cold using symptom scoring with the Jackson scale as well as by quality of life scoring with the 9-item Wisconsin Upper Respiratory Symptom Survey (WURSS-21) Quality of Life (QoL) sub-score.^14^ The preceding study also investigated whether treatment with ColdZyme in addition to optional care, when initiated at the first self-perceived prodromal symptoms could alleviate and shorten a common cold, in comparison to optional care only (“no treatment” group). The 9-item WURSS-21 QoL domain composite score, as well as eight out of nine individual QoL items, were found to be the most sensitive instrument for demonstrating that treatment with ColdZyme compared to “no treatment” significantly improved the common colds sufferers’ quality of life. The reduction in Jackson score AUC (area under the curve), where a lower value corresponds to fewer complaints and a higher value corresponds to more complaints, was likewise significantly reduced with ColdZyme treatment, although less discriminating than the 9-item WURSS-21 QoL domain. These results, together with significantly less use of optional care (paracetamol, ibuprofen, saline water nose drops or spray, antibiotics) and a quicker recovery indicated a positive effect after treatment with ColdZyme.

The aim of this present study was to evaluate the efficacy of ColdZyme for common cold, with the hypothesis that ColdZyme is superior to placebo in the treatment of naturally occurring common colds. The primary objective was to evaluate the impact on quality of life during a common cold episode, based on the primary endpoint defined as the AUC of WURSS-21 QoL composite sub-score over 8 days following start of symptoms. Day 1 for all common cold diary endpoints was defined as the first day of symptom recording when the Jackson score (sore throat, blocked nose, runny nose, cough, sneezing, headache, malaise and chilliness) in the subject’s common cold diary was at least 1 (mild = present, but not disturbing or irritating) for any symptom except headache. Additionally, the safety of ColdZyme was assessed.

## Material and methods

### Study design and participants

This trial was conducted between January and April 2019, prior to the global outbreak of SARS-CoV-2. It was a prospective, multicenter, randomized, double-blind study to evaluate the efficacy of ColdZyme for common cold using a parallel-group design that compared the investigational device to placebo in subjects with naturally occurring common colds. After approval from four local Ethics Committees (Ethikkommission der Charité, Berlin; Landesärztekammer Brandenburg, Cottbus; Sächsische Landesärztekammer, Dresden; and Landesärztekammer Baden-Württemberg, Stuttgart) and after providing written informed consent, subjects were recruited to ten health care centers in Germany.

Men and women (18-70 years) with at least three occurrences of common cold within the previous 12 months, but otherwise in good health and willing to comply with the trial procedures, were eligible and screened for enrolment at the study sites against inclusion and exclusion criteria. Women of child-bearing potential had to use appropriate contraception methods and demonstrate a negative pregnancy test at enrolment. Exclusion criteria included known allergy to the components of the investigational product, a health condition which could interfere with the results of the study or the safety of the subject, influenza vaccination within three months prior to enrolment, and regular use of products that may influence the study outcome, except for the defined optional care. Pregnancy or nursing, history of or current abuse of drugs, alcohol or medication, participation in the present study of a person living in the same household as the subject, inability to comply with study requirements according to investigator’s judgement or participation in another clinical study in the 30 days prior to enrolment and during the study, were also exclusion criteria. The inclusion and exclusion criteria were identical to the criteria in the previous trial.^14^

After enrolment and randomization, the study was conducted in an initial surveillance phase (checking daily for any onset of the common cold symptoms, documented in the diary), followed by a treatment phase initiated once the participants experienced first perception of a common cold. During the conduct of the study the number of subjects to be enrolled was raised from 600 to 701 in order to reach the required number of subjects experiencing common cold symptoms.

Prior to enrolment the trial was registered at ClincialTrials.gov, number NCT03794804. The trial was performed in compliance with the principles of the World Medical Association (Declaration of Helsinki), ICH GCP E6, German Act on Medical Devices (MPG) §23b and ISO 14155.

### Randomization and blinding

The study was conducted in a double-blind randomized manner. The subjects were randomized (1:1) at the screening visit to the ColdZyme or placebo group. Several whole blocks were allocated to each study center. The investigational products, containing ColdZyme or placebo, were identical in appearance, as well as packaging and labelling, to ensure blindness to treatment assignment for subjects and all involved site personnel.

### Interventions

All subjects received the subject diary to be used throughout the study. The diary comprised a daily question with regard to a possible onset of common cold symptoms and, after onset, specific questions to record the daily symptoms (Jackson, WURSS-21 QoL, Sore Throat Scale). Those randomized to the ColdZyme group received the investigational device, a throat spray consisting of glycerol, water, trypsin secreted from the pyloric caeca of the Atlantic cod, ethanol (<1 %), calcium chloride, trometamol and menthol (ColdZyme^®^, Enzymatica, Sweden). Subjects randomized to the placebo group received a throat spray consisting of ethanol (<1 %), menthol and water. The participants were instructed to start the treatment phase of the study when answering “Yes” to the question “Do you think you might be having the first signs of a common cold?” and simultaneously experiencing a total Jackson score of at least 1 for any symptom except headache. The treatment had to be sprayed twice (1 dose=340 µl) every second hour, up to 6 times daily.

From the first day of entering the treatment phase the subjects started to record their symptoms twice daily, in the morning and in the evening, on the Jackson scale^15^ and once daily, in the evening, on the 9-item WURSS-21 QoL domain^16^ and the Sore Throat Scale.^17,18^ All subjects were requested to answer a daily question in the evening “Do you think that you are still having a common cold?”. They also recorded the use of the investigational product and any rescue treatment and if they stayed home (away from work, school etc.) due to common cold. Entries to diaries continued for 2 days after being symptom free (as determined by answering “No” to the question above for 2 days in a row), but no longer than 10 days in total.

The Jackson score was calculated by summing the scores for the following 8□symptoms: sore throat, blocked nose, runny nose, cough and sneezing (local symptoms) as well as headache, malaise, and chilliness (systemic symptoms). Symptoms were assessed on a 4-point scale: 0□=□none, 1□=□mild, 2□=□moderate, 3□=□severe. The 9 item (items 12 to 20) QoL domain of the WURSS-21 was calculated by summing the individual scores recorded for the question “How much has your cold interfered with your ability to…”: Think clearly, sleep well, breathe easily, walk-climb stairs-exercise, accomplish daily activities, work outside the home, work inside the home, interact with others, and live your personal life^16^. Response options ranged from 0 = not at all, to 7 = severely. A lower value corresponds to an improved quality of life during illness.

Within 3 days of experiencing the first signs of common cold symptoms, the subjects presented to the investigator for a physical examination and confirmation of the common cold (Visit 2). The third and last visit to the study site for subjects having experienced a common cold had to take place by day 16 (+/-4 days) after symptom start (Visit 3). Subjects with no symptoms during the study period (who thus did not use the study treatment) only had a termination visit 16 weeks (+/-7 days) after enrolment.

Both groups were allowed to use optional care (as “rescue” treatment) comprising paracetamol (acetaminophen), maximum 2□g/day, ibuprofen, maximum 400 mg/day or saline nose drops or spray, if needed for symptom relief. Antibiotics were allowed if medically required for another ailment following the confirmation of a bacterial infection however, were not to be used for common cold.

### Outcomes

The aim of the study was to evaluate the efficacy of ColdZyme for common cold, with the study hypothesis that treatment with ColdZyme is superior to placebo in the treatment of naturally occurring common cold. Thereby, the primary objective was to evaluate the impact on quality of life during a common cold episode, based on the primary endpoint defined as the AUC of WURSS-21 QoL composite sub-score for 8 days following start of symptoms (day 1 is the first day of symptom recording).

Secondary outcome parameters included two hierarchically ordered major secondary endpoints (1 is highest ranking):

1. AUC days 1-8 composite daily severity of all symptoms within the Jackson score (mean of morning and evening) (day 1 is the first day of symptom recording)
2. Exposure to any concomitant treatment (including natural health products) that may affect common cold symptoms; immune suppressants/immune stimulants, analgesics/anti-rheumatics, anti-phlogistics, antitussives/expectorants, mouth or throat therapeutics, decongestants, antibiotics, anti-histaminergic drugs, nasal drops/spray or any medication/treatment known to affect common cold symptoms at any dose, expressed as number of days with concomitant treatment during the first 4 days for each subject (based on diary data).

Other secondary endpoints included, among others, single items of the WURSS-21 and single symptoms of the Jackson scale and their duration, day with maximum score, use of and exposure to concomitant treatment during the first 4 days, duration of the first intense phase (defined as number of days from start of treatment until Jackson score < 5, for subjects with a score ≥ 5 during the common cold episode), symptom intensity, a sore throat scale and number of days sick at home due to common cold. In addition, several safety endpoints were defined, including physical examination, vital signs and adverse events throughout the study, as well as global evaluation of tolerability by subjects and investigators at study end and assessment of device deficiencies at Visit 2 and Visit 3.

### Sample size calculation

The sample size was calculated so that the comparison of the ColdZyme group and the placebo group for the primary endpoint would have 90% power to detect a relevant clinical effect size. Based on the results of a previously conducted trial with the same investigational device in a comparable setting,^14^ n=338 subjects (with n=169 subjects per treatment group) was assessed as required for the primary confirmatory statistical analysis of the study. Of all subjects originally randomized in a 1:1 manner, only those having the specified symptoms were to actually start using the investigational product. In order to ensure the minimal number of required subjects per study group, the total number was decided to be 400 treated subjects.

### Statistical Analyses

The primary endpoint was defined as the AUC of WURSS-21 QoL composite sub-scores assessed during first 8 days of symptoms. The WURSS-21 QoL composite sub-score was calculated by summing the scores of the 9 consecutive items of WURSS-21, from item 12 (“think clearly”) to item 20 (“live your personal life”), as documented in the subject diary. The AUC was assessed by applying the trapezoidal approximation. The two-sided confirmatory null-hypothesis for the primary endpoint, the major secondary endpoints and continuous other secondary endpoints were analyzed for between-group comparison using the non-parametric two-sided Wilcoxon (Mann-Whitney U) rank-sum test, unstratified as well as stratified by study center (Van-Elteren or generalized Mann-Whitney test) to assess the treatment effect adjusted by the influence of sites. The stratified Van-Elteren test will be presented below for this multi-center trial. Multiple testing was performed without exploratory adjustment. For categorical other secondary endpoints, χ2 tests, unstratified as well as stratified by study center (Cochran-Mantel-Haenszel test), were used for between-group comparison. For continuous data, standard statistical characteristics were presented (number of subjects with non-missing data, mean, standard deviation, median, minimum and maximum and quartiles). Categorical data (nominal or ordinal) were summarized using frequency tables.

The statistical analyses were defined as confirmatory for the primary and, in a hierarchical order, the two major secondary endpoints, and exploratory for all other secondary endpoints. All inferential statistical testing was performed as two-sided. The confirmatory significance level was fixed to a type I error rate alpha of 5% (two-sided). In order to control the overall experiment type I error rate, the primary endpoint and the two major secondary endpoints were to be tested in a fixed sequence as *a-priori* ordered hypotheses.

The full analysis set (FAS) was defined in the SAP, prior to unblinding and statistical analysis, as all subjects of the all subjects treated set (AST, n=438) for which at least one of the confirmatory endpoints could be evaluated. Incorrectly randomized subjects with an unfulfilled entry criterion were excluded from the FAS (n=436) if the specific criterion violation had been measured and documented prior to randomization and, before breaking the blind, was considered to be of major importance. Analyses with respect to safety were performed for the AST. Analyses with respect to efficacy were conducted both for the FAS and the per-protocol set (PPS). Summary results are presented here for the FAS. The statistical analysis was performed using the SAS^®^ software package version 9.4 under the Microsoft Windows^®^ 10 operating system at acromion GmbH.

## Results

In the winter period of 2019, between January and April, 701 eligible subjects were recruited and randomly assigned to the ColdZyme group (n=351) or the placebo group (n=350). The last subject completed the study in May 2019. Of the 701 subjects, 438 (62.5%) subjects developed symptoms of common cold, and all 438 started study treatment for their common cold episode (n=220 in the ColdZyme group and n=218 in the placebo group). Two subjects of the AST (n=438) were excluded from the FAS (n=436), as seen in Figure 1. Baseline characteristics are described in Table 1.

**Table 1:** Baseline characteristics (Visit 1) of the full analysis set (FAS) population. Data are mean (SD); n (%).

|  | **Treatment group**<br>(n=218) | **Control group**<br>(n=218) |
| --- | --- | --- |
| Age (years) | 40.9 (14.1) | 41.7 (14.6) |
| Sex (female) | 153 (68.1 %) | 144 (66.1 %) |
| Caucasian | 215 (98.6 %) | 214 (98.2 %) |
| Systolic blood pressure (mm Hg) | 123.9 (12.8) | 123.6 (13.6) |
| Diastolic blood pressure (mm Hg) | 77.0 (8.5) | 76.7 (7.9) |
| Pulse rate (bpm) | 70.9 (9.0) | 71.1 (9.2) |

**Figure 1:**
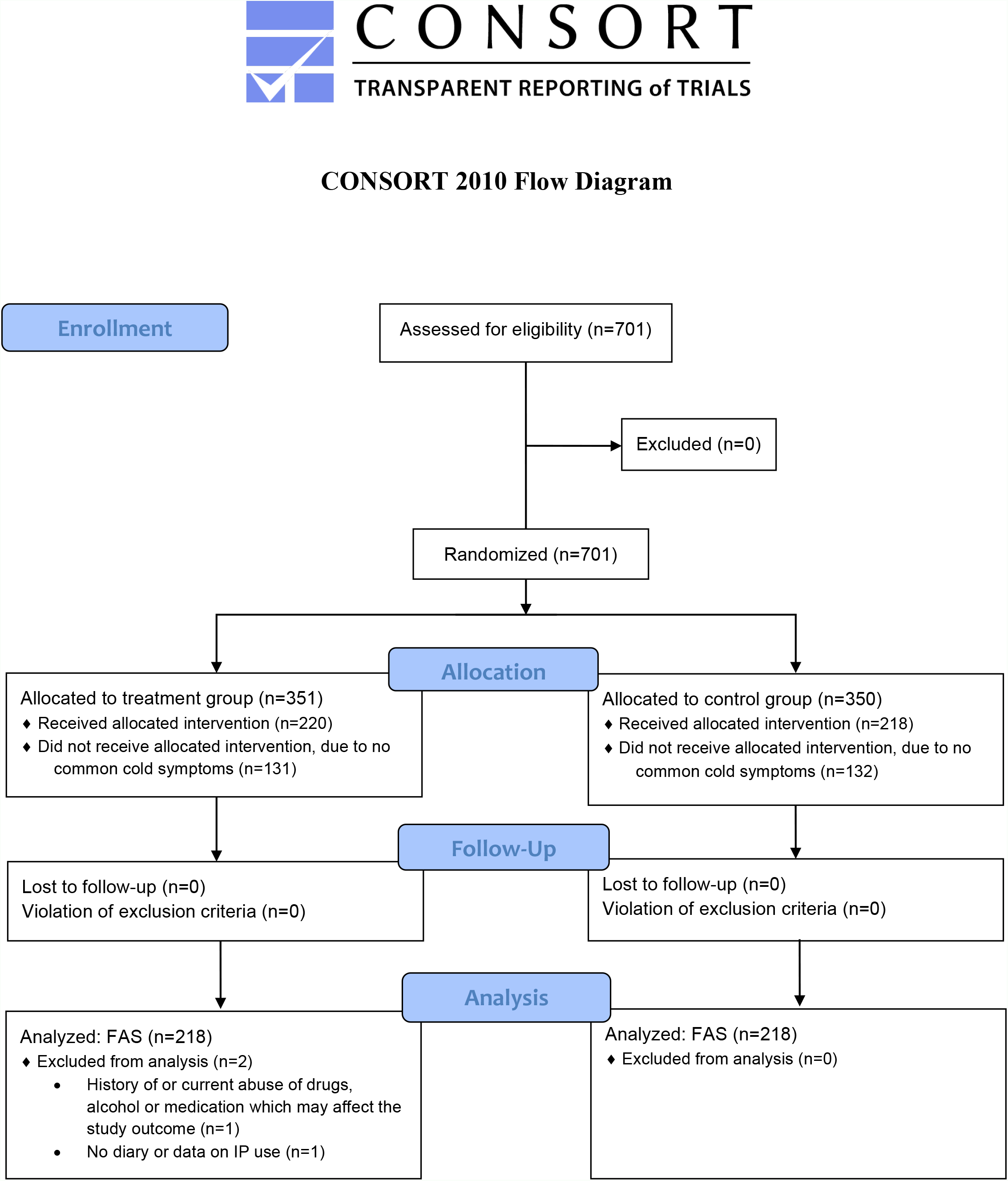
Trial profile.

Vital signs obtained at Visit 1 showed no difference between the treatment groups. The common cold started with a similar mean burden of symptoms in the two groups, with a Jackson score of 7.3 ± 3.6 (mean and standard deviation, SD) for the ColdZyme group and 7.1 ± 3.7 for the placebo group on the evening of Day 1, which was similar to the previous single-blind trial (7.5 in the treated group).^14^ The common cold was confirmed by a blinded investigator within three days from the start of a perceived common cold at Visit 2 in 98.2% of the subjects in the ColdZyme group and 99.1% in the placebo group, with no statistically significant difference.

### Efficacy

The primary endpoint was defined as the AUC for the WURSS-21 QoL composite sub-score assessed during the first 8 days of common cold symptoms (AUC_WURSS21-QoL_). No statistically significant difference was seen between the groups. The mean for the first 8 days of symptoms was 127.6 ± 85.9 in the ColdZyme group and 127.5 ± 88.3 in the placebo group (p=0.705).

As the confirmatory test for the primary efficacy endpoint failed statistical significance, both major secondary endpoints were evaluated exploratory. The first major secondary endpoint was defined as the AUC for the composite daily severity of all symptoms within the Jackson scale assessed during the first 8 days of common cold symptoms (AUC_Jackson_), based on the daily records (twice per day) for the Jackson scale items in the subjects’ common cold diary. The results were similar between the groups, 40.9 ± 24.2 in the ColdZyme group and 41.3 ± 23.4 in the placebo group (p=0.481).

The second major secondary endpoint was defined as the number of days with concomitant treatment possibly affecting common cold symptoms assessed during the first 4 days for each subject, based on the diary entries. Identification of treatments that could possibly affect common cold symptoms was made independently by an external physician. Most subjects (70.4%) had no days with treatment that might affect common cold symptoms and 14.4% of subjects had only one day. In total, 4.1% of the subjects used concomitant treatment that might affect common cold symptoms on all four days. The difference between the groups was not statistically significant (p=0.885).

Significant differences or trends for significance in favor of ColdZyme were shown for several exploratory endpoints. The mean number of days with a WURSS-21 QoL score > 0, a measure of illness duration, showed a significant difference between the groups, 6.5 ± 2.7 days (6.0 day median) in the ColdZyme group and 6.9 ± 2.6 days (7.0 day median) in the placebo group (p=0.050). Likewise, the total number of days of greater than zero on the Jackson symptom score was 6.8 ± 2.6 vs. 7.1 ± 2.6 respectively (p=0.087) with a median difference of 1.0 day (6.0 vs. 7.0) indicating the median time to recovery was 1 day sooner for the ColdZyme group. These estimates are conservative in that these outcomes require a symptom free or condition free return to baseline. These findings confirm previous observations of shortened common cold duration when using ColdZyme.^13-14,19^

The beneficial effect of ColdZyme was particularly noticeable on the fifth day of the common cold, often the pivotal day to restoring wellness. The assessment of symptom severity per Jackson score showed 11% less severe symptoms in the ColdZyme group (4.8 ± 5.0) compared to placebo (5.4 ± 4.8, p=0.066) on this day with a median difference of 1 (score 3.5 vs. 4.5). Further, 10% fewer subjects reported WURSS-21 QoL scores greater than zero in the ColdZyme group (p=0.029). Moreover, the proportion of subjects reporting to having no common cold on Day 5 was 33.5% for the ColdZyme group vs. 22.0% for the placebo group (p=0.005).

The ColdZyme group showed broadly similar scores to the control group in the remaining secondary endpoints comprising the single items and symptoms of the WURSS-21 and Jackson scales, day with maximum score, use of and exposure during the first 4 days to concomitant treatment, duration of the first intense phase, symptom intensity, sore throat scale and number of days sick at home due to common cold. At study end, subjects evaluated the global efficacy of the investigational product on a scale of “very good”, “good”, “moderate” and “poor”. A statistically significant difference between treatment groups was seen as 70.6% of subjects rated the efficacy as “very good” or “good” in the ColdZyme group as opposed to 60.1% in the placebo group (p=0.001). The investigators’ assessed the efficacy of the investigational product similarly, with ratings “very good” or “good” for 64.3% of the subjects in the ColdZyme group, as opposed to 59.2% in the placebo group (p=0.045).

### Compliance

Compliance at the start of investigational product use, defined as applying the product by the morning of the next day after the first day of a defined common cold, was similar in the two treatment groups, with 98.6% of subjects starting to use the investigational product in time as reported in the subject diary.

Evaluation of compliance to investigational product usage was performed based on the return and weighing of unused investigational product as compared to duration of the period defined for usage. A compliance of at least 80% of the correct quantity of the investigational product was seen for 265/436 (60.8%) of subjects, with a higher compliance in the ColdZyme group (64.7%) compared to the placebo group (56.9%).

### Safety

There were no relevant differences in vital signs or physical examination findings between the treatment groups. The incidence rates of TEAEs (treatment-emergent adverse events) in the ColdZyme group and the placebo group were comparable; 4.5% and 4.6% respectively. Two subjects had TEAEs with intensity “severe”: one subject in the ColdZyme group had an ileus with abdominal pain, the only serious TEAE in the study, and one subject in the placebo group had a panic disorder. Likewise, the rate of events classified as adverse device effects was similar, with 1.4% subjects of the ColdZyme group and 2.3% of subjects of the placebo group having the highest relationship to the investigational product assessed as “related”. The events comprised nausea, oral pain, pharyngeal swelling, pharyngeal hypoesthesia, dry throat and panic disorder. At the end of the study an equal number of subjects rated the tolerability of the investigational product as either “very good” or “good”, 92.7% for ColdZyme and 93.5% for placebo (p=0.135). The investigators’ rating of tolerability was “very good” or “good” for 91.9% of subjects in the ColdZyme group and 93.6% in the placebo group (p=0.129).

## Conclusion

Previous research results evaluating ColdZyme and oral mouth rinses on common cold viruses and SARS-CoV-2 infections show all have *in vitro* antiviral efficacy and when applied sufficiently onto the oral mucosa reduce viral load.^10-13,20-21^ This double-blind, placebo-controlled, randomized trial was conducted to evaluate the efficacy of ColdZyme in the treatment of naturally occurring common cold. The study followed a previous single-blind trial investigating the use of two different scales to assess the common cold. Primary and secondary endpoints were chosen based on the results of the previous study where the 9-item WURSS-21 QoL domain composite score were shown to be a sensitive instrument for demonstrating that treatment compared to no treatment significantly improves the common colds sufferers’ quality of life^14^. The reduction in Jackson symptom score was likewise significantly reduced with ColdZyme, although less discriminating than the 9-item WURSS-21 QoL domain. Those results, together with significantly less use of optional care, were indicative of a positive effect after treatment with ColdZyme.

The design of the previous and present trials was similar to several other studies assessing interventions at the first sign of self-perceived common colds illness.^8,16,22^ In order to reduce virus propagation and replication, it is essential that the tested throat spray ColdZyme is applied as early in the disease onset as possible. Since no objective signs of a common cold are present during the short prodromal phase, personal experience catching common colds makes the best predictor of imminent illness. This trial design in which ColdZyme treatment was compared to what may be considered placebo failed to find differences in its main endpoints. Limitations in the experimental design, discussed below, may account for those findings. Notwithstanding, several results suggest that duration of illness was reduced as it was in prior studies of differing design.

The previous single-blind trial determined common cold symptom durations of 6.3 and 7.1 days (treatment) and 7.1 and 8.1 days (no treatment) using two measures of duration; number of days from symptom start until the last day before answering “No” to the question “Do you think that you still have a common cold”, and number of days with a total Jackson score > zero.^14^ Though not as pronounced as in the previous ColdZyme trial, the duration of illness marking a return to wellness (as judged by the QoL subdomain returning to zero) was also shorter in the present study. The subdomain mean scores were 6.5 days and 6.9 days for the ColdZyme and placebo groups respectively, reflecting a 0.4-day shorter illness with ColdZyme; however, the median times differed by a full day (6.0 vs. 7.0; p=0.05, n=218/218). A 1-2 day reduction in a common cold episode is consistent with other treatments of common colds,^23-24^ but also how often subjects were asked to assess the presence of illness in this trial (once a day). The return to wellness was also heralded by a significant difference in the proportion of subjects on the preceding day (Day 5) who had recovered as measured by subdomain scores of zero (24.2% vs. 16.0%, p=0.029). Significant reductions in a common colds episode’s duration have also been found using other topical oral dose forms such as antiviral zinc lozenges.^25,26^ At the end of this trial more subjects (70.6%) reported a benefit with ColdZyme by rating efficacy as “very good” or “good” in the ColdZyme group as opposed to the placebo group (60.1%), while tolerability was similar (92.7% and 93.5%, respectively). Overall treatment response at end of study based on a subject’s global assessment of efficacy and tolerability reflects a broad real-world assessment.

Design limitations of this trial may have acted to reduce the ability to appreciate differences between placebo and ColdZyme treated groups affecting the primary and major secondary variables. First, the placebo spray containing menthol may have signaled a perception of efficacy in the placebo-treated subjects as sixty percent at the end of the study rated efficacy as “very good” or “good”, thereby lessening differences with active treatment. Topical menthol is well-known to act on mucosal TRPM8 sensory receptors in the oropharyngeal cavity, perceived as cooling sensations that provide an increased sensation of nasal airflow, improve physiological performance, and give a sense of reward.^27^ Menthol is an approved topical antitussive in the United States and is found in over half of consumer cough formulae surveyed.^28^ Second, the use of oral analgesics and nasal saline in the first 4 days when symptoms peak were equally high in both groups (29% of subjects), and thus may have diminished overall symptom intensity making it harder to statistically appreciate subjective differences in milder illness. Third, dosing compliance of at least 80% of the intended quantity of the investigational product, although 8% higher in the ColdZyme group than in the placebo group, was only 64.7%. Lastly, the experimental design of this trial did not include a concomitant untreated group, nor was virus presence or type confirmed.

The outcome of this study supporting a shorter duration of the common cold adds to previous findings in a randomized placebo-controlled study on healthy volunteers treated with ColdZyme, in which a lower oropharyngeal virus load and a reduced duration of common cold symptoms was shown following experimental inoculation with rhinovirus-16.^13^ Further, the study also supports findings from a study on naturally occurring upper respiratory tract infection (URTI) in competitive endurance athletes in which 123 endurance-trained, competitive athletes were randomized to control (no treatment, n=61) or ColdZyme (n=62) for a 3-month study period.^19^ They recorded daily training and illness symptoms during the study period. The study found that ColdZyme was able to reduce the duration of the common cold, symptom severity ratings, and the associated number of missed training days. Lastly, the safety and tolerability of ColdZyme shown in previous trials^13-14,19^ has now been confirmed in a large study population.

## Data Availability

Individual participant data, after having been anonymized, will be made available, along with the study protocol, statistical analysis plan and informed consent form template. Data will be available beginning 3 months and ending 5 years after publication of the article to researchers whose proposed use of the data is approved by the study sponsor. Proposals should be directed to the corresponding author and requesters will need to sign a data access agreement.

## Acknowledgements

The authors would like to thank analyze & realize GmbH, Prof. Ralf Uebelhack and participating investigators and sites for their invaluable contribution to the conduct of the trial.

